# Association Between Residential Greenness and Preeclampsia in Japan: The TMM BirThree Cohort Study

**DOI:** 10.1101/2025.05.13.25327565

**Authors:** Hisashi Ohseto, Ami Uematsu, Mami Ishikuro, Zheng Xian, Yuta Takahashi, Masatsugu Orui, Keiko Murakami, Aoi Noda, Genki Shinoda, Geng Chen, Noriyuki Iwama, Masahiro Kikuya, Hirohito Metoki, Atsushi Hozawa, Taku Obara, Tomoki Nakaya, Shinichi Kuriyama

## Abstract

**Background:** Preeclampsia (PE), characterized by hypertension and organ dysfunction during pregnancy, is a leading cause of maternal and fetal mortality. Residential greenness has been reported to be negatively associated with a broad range of health outcomes, such as mental illness and cardiovascular disease. However, evidence on the association between residential greenness and PE remains limited, particularly among non-European populations.

**Objective:** This study aimed to investigate the association between residential greenness and PE among pregnant women in Japan, considering urbanization status and the timing of PE onset.

**Methods:** This study included 21,816 pregnant women from the Tohoku Medical Megabank Project Birth and Three-Generation Cohort Study, conducted in Miyagi Prefecture, Japan. Residential greenness was assessed using the Normalized Difference Vegetation Index (NDVI) values calculated from the center of each participant’s postal code area. PE was identified using a rule-based phenotyping algorithm applied to medical records. Logistic regression analyses were conducted to estimate odds ratios (ORs) and confidence intervals (CIs), adjusting for geographical variables, such as air pollution, urbanization status, area deprivation index, and individual-level confounders.

**Results:** Moderate NDVI levels within a 200 m buffer were associated with a lower incidence of PE than low NDVI levels (OR: 0.79 [95% CI: 0.63–1.00]). High NDVI levels also suggested a negative association, but the results were not statistically significant (OR: 0.85 [95% CI: 0.64–1.14]), and no clear trend was observed (P for trend = 0.235). After adjusting for potential mediators, including psychological distress and physical activity, estimated values remained unchanged, but associations lost statistical significance. This association was primarily observed in non-urban areas and in late-onset PE.

**Conclusion:** Moderate residential greenness was significantly associated with a lower incidence of PE compared to low residential greenness. These results suggest that moderate residential greenery is worth considering when choosing where to live during pregnancy.

## Introduction

Preeclampsia (PE), a subtype of hypertensive disorders of pregnancy (HDP), is characterized by hypertension and organ dysfunction after 20 weeks of gestation. Despite its relatively modest prevalence—approximately 3.4% and 2.7% in pregnant women in the USA^1^ and Japan^2^ — PE is one of the leading causes of maternal and fetal mortality.^3–6^ The pathophysiology of PE involves systemic inflammation and abnormal immune responses caused by placental dysfunction, distinguishing it from essential hypertension.^5,7,8^ Therefore, while several risk factors are shared with essential hypertension, such as advanced age, obesity, and a family history of hypertension,^9^ PE also has unique risk factors, including nulliparity, short inter-birth intervals, and changes in sexual partners.^10,11^ A key question in maternal and child health is whether insights from essential hypertension research can be extended to PE prevention or management.

Residential greenness, an indicator of vegetation abundance in the surrounding environment, is typically quantified using measures such as the Normalized Difference Vegetation Index (NDVI) derived from satellite imagery or green space proportions based on land-use data.^12^ Previous studies have shown that greenness has protective effects on a broad range of health outcomes such as mental illnesses (e.g., depression and anxiety), cardiovascular diseases, and all-cause mortality.^13–15^ Regarding blood pressure (BP), studies have reported antihypertensive effects of greenness surrounding residential areas^16,17^ and schools,^18^ with these effects observed regardless of age or sex. Proposed mechanisms linking greenness exposure to health benefits include direct effects such as a reduction in air pollution^19^ and indirect effects such as stress reduction and promotion of physical activity.^20^ Additionally, exposure to natural environments may alter immune responses and the human microbiome, potentially influencing health outcomes.^21–23^ Given that the pathology of PE involves inflammation, abnormal immune responses, and a predisposition to high BP, greenness may exert a protective effect against PE, potentially through its anti-inflammatory and immunomodulatory properties.^5,7,8^ However, results from six previous studies,^24–29^ including ecological studies,^29^ have been inconsistent, only the most recent study reported a protective association between greenness and PE,^28^ necessitating further research and validation. As all these studies were conducted in the United States, validating the results in Japan, where vegetation and geographical characteristics differ, would provide important public health insights for Japan and, if reproducible, would demonstrate greater external validity beyond regional and ethnic differences.

Therefore, this study investigated the association between residential greenness and PE in a prospective cohort of 21,816 Japanese pregnant women. Secondary analyses included stratification by urbanization status and outcome-specific analyses of other HDP subtypes and timing of PE onset.

## Methods

### Participants

This prospective cohort study recruited 23,406 pregnant women in Miyagi Prefecture, Japan, from 2013 to 2017 as part of the Tohoku Medical Megabank Project Birth and Three-Generation (TMM BirThree) Cohort Study,^30,31^ which involved approximately 50 obstetric clinics and hospitals. Participants who withdrew consent and those with missing data on pregnancy status, multiple births, invalid address data, and missing PE or other HDP statuses were excluded, leaving 21,816 participants for the main analysis (Figure 1). Ethical approval was obtained from the Ethics Committee of the Tohoku Medical Megabank Organization (2013-1-103-1), and all participants provided written informed consent for study participation.

**Figure 1.**
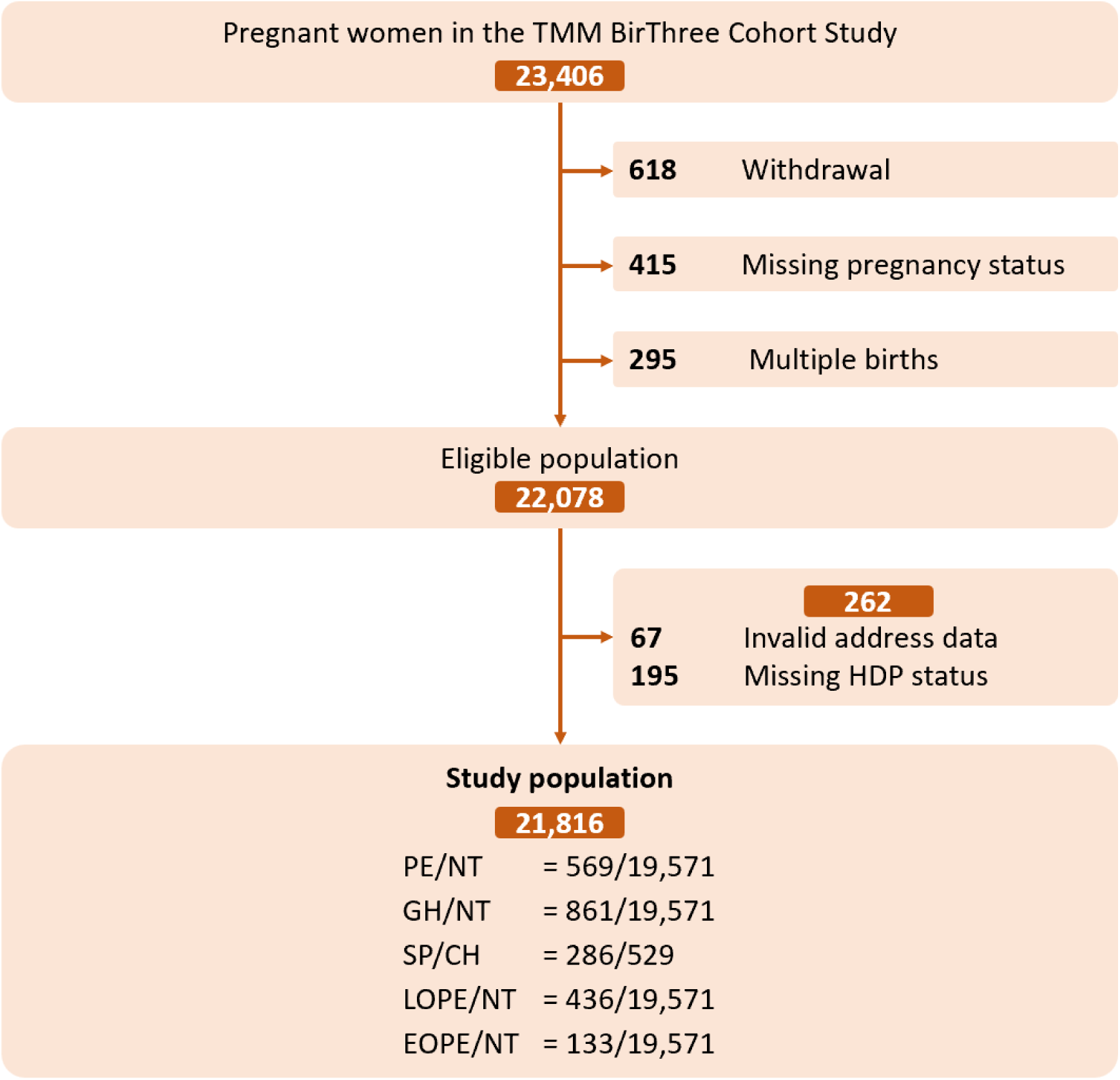
Flow chart of the present study. A total of 22,078 participants fulfilled the eligibility criteria and the main analysis was conducted on 21,816 participants with complete addresses and HDP status data. TMM BirThree Cohort Study, Tohoku Medical Megabank Project Birth and Three-Generation Cohort Study; HDP, hypertensive disorders of pregnancy; PE, preeclampsia; NT, normotension, GH, gestational hypertension; SP, superimposed preeclampsia; CH, chronic hypertension; EOPE, early-onset preeclampsia; LOPE, late-onset preeclampsia.

### Geographic Variables

We assessed residential greenness using the NDVI, derived from Landsat 8 satellite imagery, with a spatial resolution of 30 m via the Google Earth Engine (GEE).^32,33^ NDVI was derived from the near-infrared and red spectral bands, ranging from −1 to 1, where negative values indicate blue spaces (e.g., water bodies and ice surfaces), and positive values represent increasing vegetation density from bare soil (near 0), through grassland and shrubland, to dense forest. Cloud and cloud shadow contamination were removed using masking algorithms and monthly composites were generated to minimize data gaps. We selected all remote sensing images from January, April, July, and October 2015 and calculated the annual mean NDVI to minimize seasonal variations in greenness. This period represents the midpoint of the recruitment phase (2013–2017) and was used as a representative measure, assuming minimal short-term changes in residential greenness.^34,35^ Area-based mean NDVI values were calculated within buffers of 200, 500, and 1000 m radii around the population-weighted centroid coordinates of each residential postal code area reported at recruitment, aiming to evaluate greenness exposure at varying spatial scales and identify the scale most relevant to PE. The minimum buffer of 200 m was determined by the spatial resolution limit of postal code-based data, while the 500 m and 1000 m buffers were selected based on commonly used buffer sizes in NDVI studies targeting pregnant women.^36^ Based on the tertiles of mean NDVI values within each buffer, greenness exposure was classified into three levels: low, moderate, and high.

In addition to NDVI, we included the following geographic variables as covariates. The urbanization level of the participants’ places of residence was categorized into six groups (metropolitan areas, large cities, accessible small towns, remote small towns, accessible rural settlements, and remote rural settlements) and, in this study, reclassified into a binary variable: ‘urban’ (Sendai City, the capital and largest city of Miyagi Prefecture) and ‘non-urban’ (all other areas).^37^ Air pollution was assessed by predicting mean annual concentrations of NO_2_ and PM_2.5_ in 2015 using universal kriging models, with predictor variables including traffic intensity, population, land use, elevation, and geographic coordinates. The 10-fold cross-validated R^2^ was 0.67 for NO_2_ and 0.63 for PM_2.5_. Areal deprivation index (ADI) was calculated using Japan’s 2015 Population Census.^38^ ADI is a composite indicator defined as the weighted sum of the proportions of socially and economically disadvantaged groups, including older couple households, older single households, rental households, single-mother households, sales and service workers, agricultural workers, blue-collar workers, and unemployed individuals. The postal code-level ADI was estimated by proportionally allocating the census-level ADI values to the respective areas.

### Outcomes

The primary outcome measure was the presence of PE. Secondary outcomes included other HDP subtypes—gestational hypertension (GH), superimposed PE (SP), and timing-based classification of PE into early-onset (EOPE) and late-onset (LOPE). HDP subtypes were identified through an automated, rule-based phenotyping algorithm that utilized medical records detailing hypertensive disease history, BP, proteinuria, timing of onset, and PE-related clinical conditions.^39,40^ The accuracy of this algorithm was confirmed through physician validation.^39^ Hypertension was defined as a systolic BP (SBP) of 140 mmHg or higher, a diastolic BP (DBP) of 90 mmHg or higher, or both, measured during one antenatal checkup. GH was characterized by the absence of pre-existing chronic hypertension (CH) and the development of hypertension after 20 weeks of gestation without proteinuria. A diagnosis of PE required the absence of pre-existing CH and early pregnancy proteinuria, along with the presence of hypertension and proteinuria (≥1+ on dipstick tests) after 20 weeks of gestation. SP referred to pre-existing CH before 20 weeks of gestation accompanied by proteinuria or the emergence of PE-related conditions after 20 weeks. Additionally, PE was categorized as EOPE if it developed before 34 weeks of gestation, and as LOPE otherwise. Further details are provided elsewhere.^39^

### Covariates

Covariates included advanced maternal age at conception (<35 or ≥35 years), pre-pregnancy overweight (body mass index <25 or ≥25 kg/m^2^), maternal educational attainment (high school or lower, junior or vocational college, or university or higher), annual household income (<4, 4 –6, or ≥6 million Japanese Yen (JPY)/year), medical history of diabetes mellitus (presence or absence), medical history of systemic lupus erythematosus (presence or absence), family history of HDP (presence or absence), alcohol consumption during early pregnancy (presence or absence), smoking during early pregnancy (presence or absence), moderate-to-vigorous physical activity during pregnancy (≥150 minutes/week or not), psychological distress during pregnancy (presence or absence), social isolation during pregnancy (presence or absence), parity (nulliparous, parous with previous HDP, or parous with no previous HDP), conception via in vitro fertilization (IVF, presence or absence), offspring sex (male or female), and season of conception (winter: December–February, spring: March–May, summer: June– August, fall: September–November).^36,40–42^ Maternal age at conception, pre-pregnancy overweight, parity, offspring sex, and season of conception were obtained from medical records, while the remaining variables were self-reported. Psychological distress was assessed using the Kessler Psychological Distress Scale (K6),^43^ with a score of five or higher indicating psychological distress. Social isolation was assessed using the Lubben Social Network Scale (LSNS-6),^44^ with a score of 11 or lower indicating social isolation.

### Statistical Analysis

Geographic variables, covariates, and the incidence of PE and other HDP subtypes were compared across tertile-based mean NDVI levels within a 200 m buffer, which was used as the representative scale for descriptive analyses as it reflects proximal residential greenness and is supported by a previous study showing stronger associations with PE at smaller buffer sizes.^28^ Continuous variables were analyzed using analysis of variance, while categorical variables were analyzed using the Chi-squared test. We also conducted natural cubic spline analyses to explore potential non-linear associations between mean NDVI values within a 200 m buffer and the incidence of PE. The spline models used three degrees of freedom and were stratified by urbanization status (urban and non-urban). Participants with NDVI values outside ±3 standard deviations within each stratum were excluded from the spline analysis to minimize the influence of extreme values.

The associations between tertile-based mean NDVI levels within 200, 500, and 1000 m buffers and PE were investigated using multiple logistic regression analysis, with low mean NDVI levels as the reference, and estimates were obtained as odds ratios (ORs) with 95% confidence intervals (CIs). Four regression models were developed to evaluate these associations by progressively incorporating additional covariates. Model 1 was unadjusted and served as the crude model. Model 2 included adjustments for geographical variables (PM_2.5_, NO_2_, urbanization status, and ADI). Model 3 incorporated adjustments for geographical variables and confounders (maternal age, family history of HDP, medical history of diabetes mellitus and systemic lupus erythematosus, educational attainment, household income, alcohol consumption, smoking, parity, IVF, offspring sex, and season of conception). Model 4 was further adjusted for potential mediators (pre-pregnancy overweight, physical activity, psychological distress, and social isolation) to ensure comparability with the results of previous studies. Missing covariate data were imputed using multiple imputations by chain equations with 50 iterations.^45^

For the secondary analysis, we conducted two additional analyses. First, we examined the association between mean NDVI levels and PE, stratified by urbanization status. Second, we analyzed the association between mean NDVI levels, other HDP subtypes, and timing of PE onset. All secondary analyses, consistent with Model 4 of the main analysis, were adjusted for all covariates and geographic variables.

Participants were restricted to those without pre-existing CH when the outcomes were PE or GH, and to those with pre-existing CH when the outcome was SP (Figure 1). Additionally, to ensure that the control group consisted solely of normotensive pregnancies, participants with PE and GH were mutually excluded in each respective analysis. Similarly, participants with EOPE and LOPE were mutually excluded when analyzing these subtypes.

Statistical significance was defined as a p-value < 0.05. All analyses were performed using R software (version 4.1.2).

## Result

Table 1 shows the baseline characteristics of the study population by tertile-based mean NDVI levels within a 200 m buffer. The NDVI distribution ranged from 0.040 to 0.381, and the tertiles were defined as follows: low (<0.094), moderate (0.094–<0.128), and high (≥0.128). These values were relatively low compared to previous studies,^24,26^ likely due to the use of annual average NDVI, including winter months and the unique residential environment in Japan. Among the 21,816 participants, 7,272 (33.3%), 7,272 (33.3%), and 7,272 (33.3%) had high, moderate, and low NDVI levels, respectively (Table 1). Participants with low NDVI levels tended to be older, have lower rates of overweight, smoking, alcohol consumption, and physical activity engagement; have higher levels of education and income; report lower psychological distress, be socially isolated, be nulliparous, and conceive via IVF (Table 1). Participants with low NDVI levels also tended to live in urban areas, were exposed to higher levels of NO_2_ and PM_2.5_, and resided in less-deprived areas (Table 1). Among participants with high, moderate, and low NDVI levels, 213 (2.9%), 172 (2.4%), and 184 (2.5%), respectively, developed PE (Table 2). In the spline analysis, a non-linear relationship was observed between NDVI and the incidence of PE, with a U-shaped relationship showing the lowest incidence around NDVI values of approximately 0.12 to 0.17 (Figure 2). Overall, the incidence was slightly higher in urban areas compared to non-urban areas. A possible decrease in PE incidence was also observed in urban areas within the lower NDVI range (<0.08); however, the wide 95% CI in this range limited interpretability.

**Figure 2.**
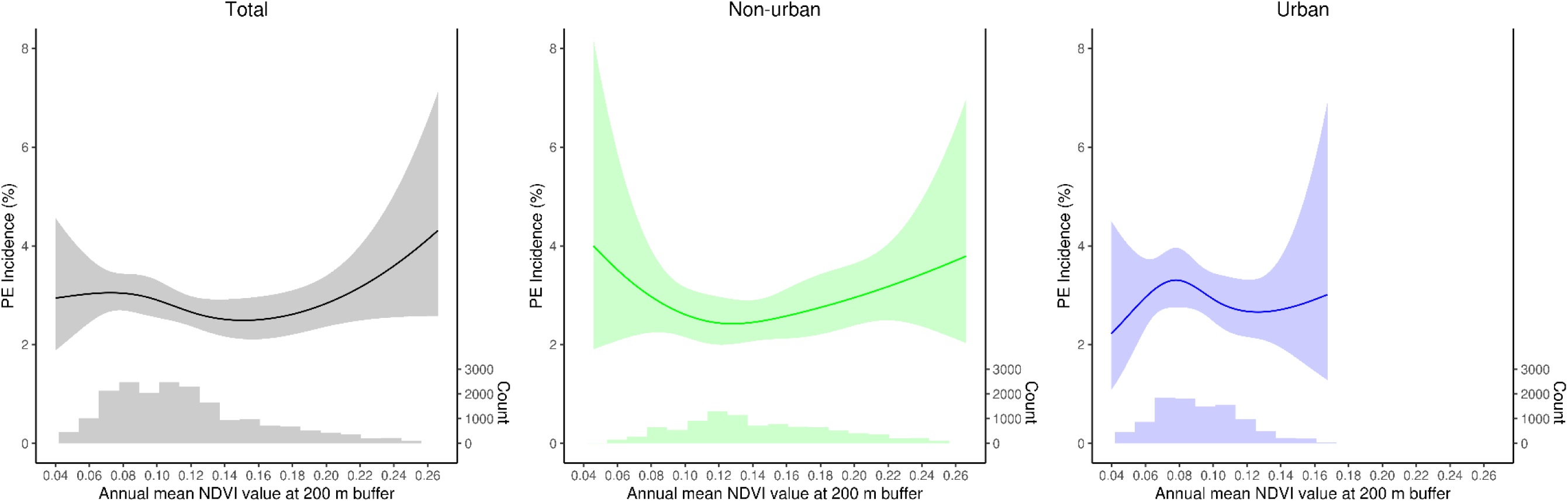
Spline analysis of the association between residential greenness and PE incidence at a 200 m buffer. Natural cubic spline models with three degrees of freedom were used to assess the non-linear association between mean NDVI values within a 200 m buffer and PE incidence, stratified by urbanization status (urban and non-urban). PE, preeclampsia; NDVI, Normalized Difference Vegetation Index.

**Table 1.**
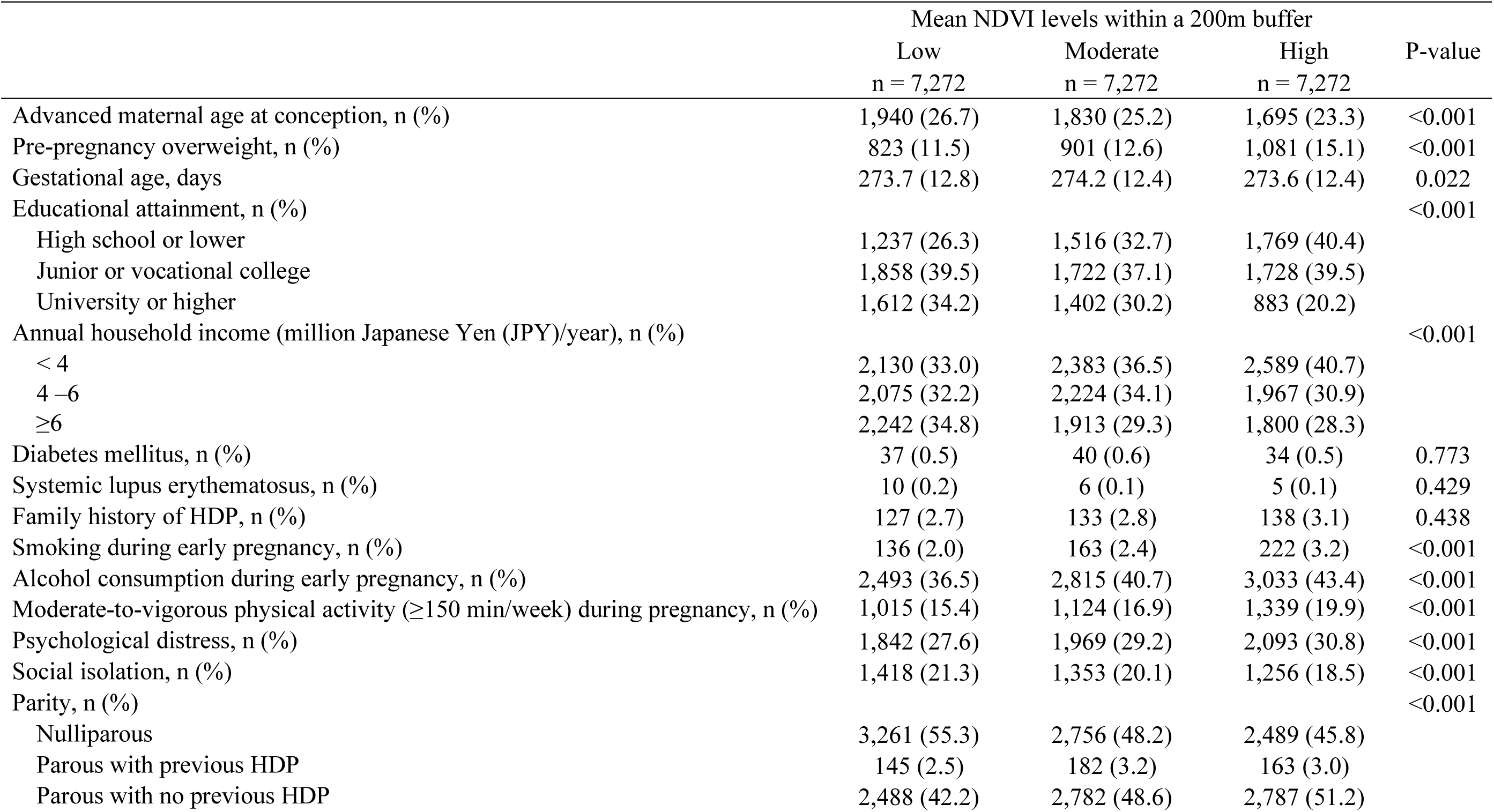
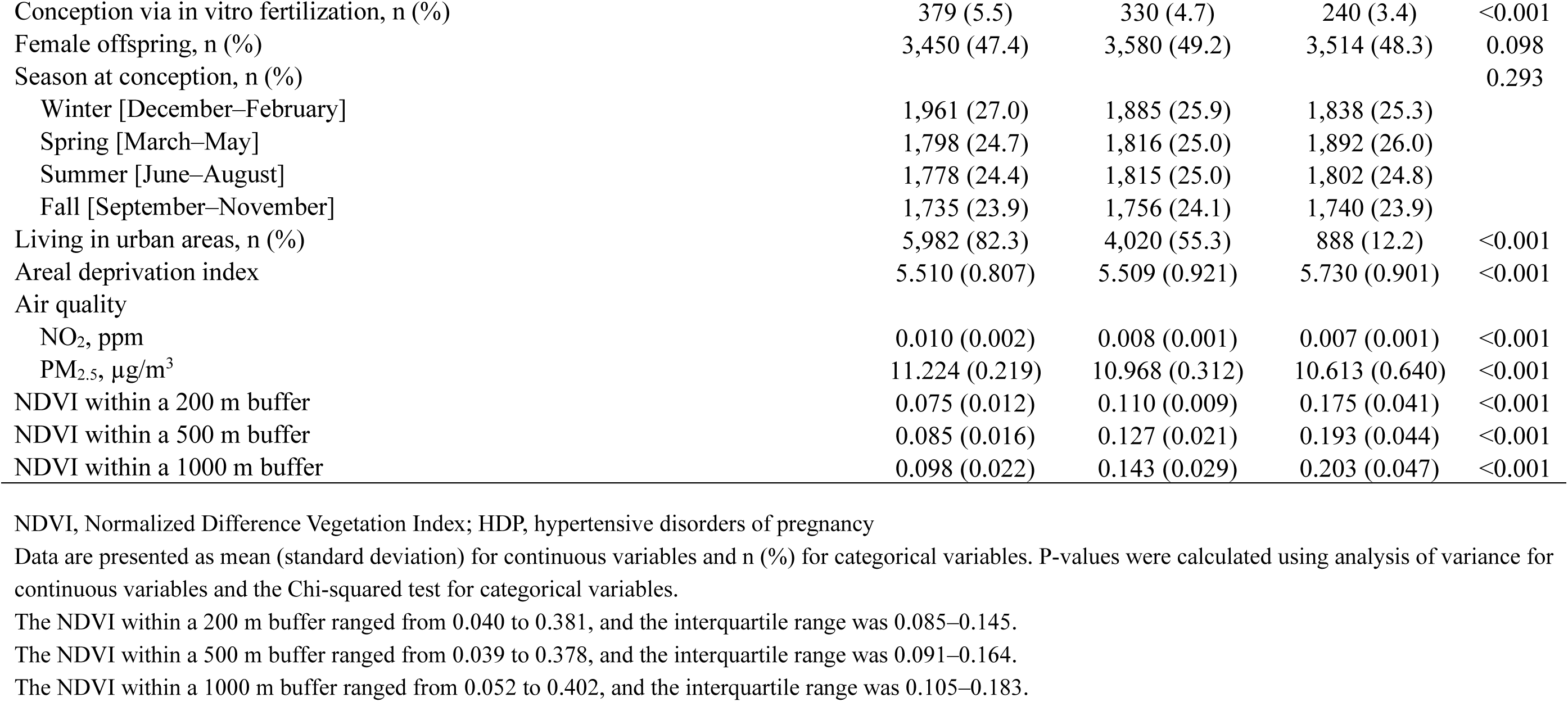
Baseline characteristics in the study population by tertiles of mean NDVI level within a 200m buffer.

**Table 2.** Incidence of outcomes in the study population by tertiles of mean NDVI level within a 200m buffer.

|  | Mean NDVI level within a 200m buffer |  |  | P-value |
| --- | --- | --- | --- | --- |
|  | Low<br>n = 7,272 | Moderate<br>n = 7,272 | High<br>n = 7,272 |  |
| HDP subtypes, n (%) |  |  |  | 0.238 |
| Normotension | 6,473 (89.0) | 6,539 (89.9) | 6,559 (90.2) |  |
| Chronic hypertension | 184 (2.5) | 186 (2.6) | 159 (2.2) |  |
| Gestational hypertension | 295 (4.1) | 284 (3.9) | 282 (3.9) |  |
| Preeclampsia | 213 (2.9) | 172 (2.4) | 184 (2.5) |  |
| Superimposed preeclampsia | 107 (1.5) | 91 (1.3) | 88 (1.2) |  |
| Onset timing of preeclampsia, n (%) |  |  |  | 0.061 |
| Early onset preeclampsia | 49 (0.7) | 49 (0.7) | 35 (0.5) |  |
| Late onset preeclampsia | 164 (2.3) | 123 (1.7) | 149 (2.0) |  |

NDVI, Normalized Difference Vegetation Index; HDP, hypertensive disorders of pregnancy
Data are presented as n (%). P-values were calculated using the Chi-squared test.

In the logistic regression analyses (Table 3), moderate NDVI levels within a 200 m buffer were negatively associated with PE onset compared to low NDVI levels, with ORs of 0.80 [95% CI: 0.65–0.98], 0.78 [95% CI: 0.62–0.98], and 0.79 [95% CI: 0.63–1.00] in Models 1, 2, and 3, respectively. High NDVI levels were also associated with a lower incidence of PE, although the results did not reach statistical significance (ORs of 0.85 [95% CI: 0.70–1.04], 0.82 [95% CI: 0.62–1.10], and 0.85 [95% CI: 0.64–1.14] in Models 1, 2, and 3, respectively), and no clear trend was observed. Although statistical significance was not attained in Model 4, the estimated values remained largely unchanged. As buffer size increased, the strength of the association decreased (Table 3). Since the ORs showed consistent values across the four models, we conducted a secondary analysis using only Model 4.

**Table 3.** The association between mean NDVI levels and PE.

| Buffer | Mean NDVI levels | Model 1 |  |  | Model 2 |  |  | Model 3 |  |  | Model 4 |  |  |
| --- | --- | --- | --- | --- | --- | --- | --- | --- | --- | --- | --- | --- | --- |
|  |  | OR [95% CI] | P-value | P for trend | OR [95% CI] | P-value | P for trend | OR [95% CI] | P-value | P for trend | OR [95% CI] | P-value | P for trend |
| 200m | Low | Reference |  | 0.111 | Reference |  | 0.147 | Reference |  | 0.235 | Reference |  | 0.230 |
|  | Moderate | **0.80 [0.65–0.98]** | **0.031** |  | **0.78 [0.62–0.98]** | **0.030** |  | **0.79 [0.63–1.00]** | **0.048** |  | 0.80 [0.63–1.00] | 0.051 |  |
|  | High | 0.85 [0.70–1.04] | 0.118 |  | 0.82 [0.62–1.10] | 0.182 |  | 0.85 [0.64–1.14] | 0.286 |  | 0.85 [0.63–1.14] | 0.277 |  |
| 500m | Low | Reference |  | 0.328 | Reference |  | 0.540 | Reference |  | 0.599 | Reference |  | 0.564 |
|  | Moderate | 0.88 [0.72 to 1.08] | 0.228 |  | 0.89 [0.71 to 1.11] | 0.297 |  | 0.89 [0.71 to 1.12] | 0.334 |  | 0.90 [0.71 to 1.13] | 0.350 |  |
|  | High | 0.90 [0.74 to 1.11] | 0.332 |  | 0.92 [0.68 to 1.24] | 0.590 |  | 0.93 [0.69 to 1.27] | 0.653 |  | 0.92 [0.68 to 1.25] | 0.611 |  |
| 1000m | Low | Reference |  | 0.795 | Reference |  | 0.632 | Reference |  | 0.684 | Reference |  | 0.700 |
|  | Moderate | 1.02 [0.83 to 1.25] | 0.875 |  | 1.06 [0.84 to 1.33] | 0.641 |  | 1.06 [0.84 to 1.35] | 0.604 |  | 1.06 [0.84 to 1.35] | 0.601 |  |
|  | High | 0.97 [0.79 to 1.19] | 0.795 |  | 1.08 [0.79 to 1.48] | 0.645 |  | 1.06 [0.77 to 1.47] | 0.707 |  | 1.06 [0.77 to 1.46] | 0.725 |  |

NDVI, Normalized Difference Vegetation Index; PE, preeclampsia; OR, odds ratio; CI, confidence interval.
ORs and 95% CIs were estimated using logistic regression analyses with low NDVI levels as references. Model 1 was unadjusted. Model 2 was adjusted for geographical variables (PM<sub>2.5</sub>, NO<sub>2</sub>, urbanization status, and the area deprivation index). Model 3 was adjusted for geographical variables and confounders (maternal age, family history, medical history, educational attainment, household income, alcohol consumption, smoking, parity, use of in vitro fertilization, offspring sex, and season of conception). Model 4 was additionally adjusted for potential mediators (pre-pregnancy overweight, physical activity, psychological distress, and social isolation).

When stratified by urbanization status, a negative association between NDVI levels and PE incidence was observed only among participants living in non-urban areas (Table 4). In non-urban areas, compared to low NDVI levels within a 200 m buffer, the ORs for PE were 0.57 [95% CI: 0.38–0.87] and 0.75 [95% CI: 0.51–1.11] for moderate and high NDVI levels, respectively. In contrast, no significant associations were observed among participants in urban areas, with ORs of 0.89 [95% CI: 0.66–1.20] and 0.69 [95% CI: 0.40–1.20] for moderate and high NDVI levels, respectively. Regarding the other HDP subtypes, the associations were not significant (Table 5). Moderate NDVI levels were negatively associated with LOPE, with an OR of 0.70 [95% CI: 0.53–0.91], but not with EOPE, with an OR of 1.21 [95% CI: 0.77–1.91].

**Table 4.** The association between mean NDVI levels and PE by urbanization status.

| Buffer | Mean<br>NDVI<br>levels | Living in non-urban areas<br>(n = 10926) |  |  | Living in urban areas<br>(n = 10890) |  |  |
| --- | --- | --- | --- | --- | --- | --- | --- |
|  |  | OR [95% CI] | P-value | P for<br>trend | OR [95% CI] | P-value | P for<br>trend |
| 200m | Low | Reference |  | 0.532 | Reference |  | 0.198 |
|  | Moderate | **0.57 [0.38–0.87]** | **0.009** |  | 0.89 [0.66–1.20] | 0.455 |  |
|  | High | 0.75 [0.51–1.11] | 0.150 |  | 0.69 [0.40–1.20] | 0.188 |  |
| 500m | Low | Reference |  | 0.381 | Reference |  | 0.865 |
|  | Moderate | **0.62 [0.41–0.93]** | **0.022** |  | 1.05 [0.78–1.42] | 0.739 |  |
|  | High | 0.74 [0.49–1.12] | 0.151 |  | 0.89 [0.53–1.49] | 0.652 |  |
| 1000m | Low | Reference |  | 0.686 | Reference |  | 0.415 |
|  | Moderate | 0.90 [0.59–1.38] | 0.632 |  | 1.18 [0.87–1.62] | 0.290 |  |
|  | High | 0.90 [0.58–1.39] | 0.627 |  | 1.14 [0.65–2.01] | 0.652 |  |

NDVI, Normalized Difference Vegetation Index; PE, preeclampsia; OR, odds ratio; CI, confidence interval. ORs and 95% CIs were estimated using logistic regression analyses with low NDVI levels as references. All results were adjusted for geographic variables (PM<sub>2.5</sub>, NO<sub>2</sub>, urbanization status, and the area deprivation index) and individual-level factors (maternal age, family history, medical history, educational attainment, household income, alcohol consumption, smoking, parity, use of in vitro fertilization, offspring sex, season of conception, pre-pregnancy overweight, physical activity, psychological distress, and social isolation).

**Table 5.** The association between mean NDVI levels and GH, SP, EOPE, and LOPE.

| Buffer | Mean<br>NDVI<br>levels | GH |  |  | SP |  |  | EOPE |  |  | LOPE |  |  |
| --- | --- | --- | --- | --- | --- | --- | --- | --- | --- | --- | --- | --- | --- |
|  |  | OR [95% CI] | P-value | P for<br>trend | OR [95% CI] | P-value | P for<br>trend | OR [95% CI] | P-value | P for<br>trend | OR [95% CI] | P-value | P for<br>trend |
| 200m | Low | Reference |  | 0.838 | Reference |  | 0.572 | Reference |  | 0.817 | Reference |  | 0.141 |
|  | Moderate | 0.99 [0.82–1.19] | 0.889 |  | 0.83 [0.55–1.24] | 0.358 |  | 1.21 [0.77–1.91] | 0.407 |  | **0.70 [0.53–0.91]** | **0.007** |  |
|  | High | 0.98 [0.77–1.24] | 0.838 |  | 0.87 [0.51–1.49] | 0.612 |  | 1.04 [0.55–1.95] | 0.902 |  | 0.80 [0.57–1.11] | 0.183 |  |
| 500m | Low | Reference |  | 0.221 | Reference |  | 0.532 | Reference |  | 0.363 | Reference |  | 0.269 |
|  | Moderate | 0.97 [0.81–1.17] | 0.746 |  | 0.79 [0.52–1.18] | 0.247 |  | 1.42 [0.90–2.24] | 0.137 |  | 0.78 [0.60–1.01] | 0.063 |  |
|  | High | 0.85 [0.66–1.09] | 0.205 |  | 0.86 [0.49–1.53] | 0.613 |  | 1.28 [0.67–2.44] | 0.454 |  | 0.84 [0.60–1.19] | 0.326 |  |
| 1000m | Low | Reference |  | 0.675 | Reference |  | 0.806 | Reference |  | 0.227 | Reference |  | 0.854 |
|  | Moderate | 1.02 [0.85–1.24] | 0.805 |  | 0.80 [0.52–1.22] | 0.299 |  | 1.42 [0.89–2.28] | 0.142 |  | 0.97 [0.74–1.27] | 0.840 |  |
|  | High | 0.94 [0.72–1.22] | 0.633 |  | 0.96 [0.53–1.75] | 0.901 |  | 1.45 [0.73–2.88] | 0.282 |  | 0.97 [0.67–1.39] | 0.861 |  |

NDVI, Normalized Difference Vegetation Index; GH, gestational hypertension; SP, superimposed preeclampsia; EOPE, early-onset preeclampsia; LOPE, late-onset preeclampsia; OR, odds ratio; CI, confidence interval.
ORs and 95% CIs were estimated using logistic regression analyses with low NDVI levels as references. All results were adjusted for geographic variables (PM<sub>2.5</sub>, NO<sub>2</sub>, urbanization status, and area deprivation index) and individual-level factors (maternal age, family history, medical history, educational attainment, household income, alcohol consumption, smoking, parity, use of in vitro fertilization, offspring sex, season of conception, pre-pregnancy overweight, physical activity, psychological distress, and social isolation).

## Discussion

### Summary

A nonlinear relationship between NDVI and PE incidence was observed in the spline analysis. Moderate NDVI levels within a 200 m buffer were associated with lower PE incidence than low NDVI levels. Although high NDVI also showed a negative association, the results did not reach statistical significance, and no clear trend was observed. In the secondary analysis, a negative association between NDVI and PE incidence was observed, primarily in non-urban areas. No association with other HDP subtypes was observed. Regarding the onset timing of PE, moderate NDVI levels within a 200 m buffer were associated with lower LOPE incidence.

### Our Results in the Context of Previous Studies

Six previous studies investigated the relationship between residential greenness and PE, including five observational studies^24–28^ and one ecological study.^29^ All of these studies were conducted in the United States and targeted relatively urbanized areas. While there are similarities with the present study, significant differences in vegetation, regional environments, ethnicity, and lifestyles between Japan and the United States necessitate careful consideration when making comparisons. This study is the only evidence to date suggesting a potential impact of residential greenness on the incidence of PE among the Japanese populations.

The results of previous studies have been inconsistent. Three studies found no association between residential greenness and PE,^26,27,29^ two reported associations only with SP,^24,25^ and one identified a significant association with PE.^28^ The earliest study,^27^ published in 2013, targeted over 80,000 pregnant women who gave birth in Southern California between 1997 and 2006. This study evaluated residential greenness within 50–150 m buffers around residences using NDVI but found no significant association with PE. Conversely, the most recent study,^28^ published in 2023, examined 1,943 pregnant women who gave birth in Philadelphia between 2013 and 2016. This study evaluated residential greenness within 100 m and 500 m buffers using tree canopy cover and identified a significant negative association with PE only within the 100 m buffer.

Regarding the methods of greenness assessment, four observational studies used NDVI,^24–27^ while one used tree canopy cover.^28^ Among these, significant associations with PE were observed only in the study using tree canopy cover,^28^ which, unlike NDVI, which reflects greenness from various types of vegetation, specifically represents tree-related greenery.

Additionally, buffer sizes ranging from 50 m to 500 m have been employed in previous studies, and the results were not necessarily consistent. For example, SP was significantly associated with residential greenness within a 500 m buffer,^24,25^ whereas PE showed a significant association within a 100 m buffer.^28^ In this study, the association became stronger as the buffer size decreased from 1000 m to 500 m and 200 m. These findings suggest that a buffer size of approximately 100–500 m may be most effective for capturing the impact of residential greenness on pregnant women. This pattern may be explained by individuals’ tendencies to engage in activities closer to their residences.

### Non-Linear Relationships

The relationship between greenness exposure and health outcomes is often nonlinear. For instance, a previous study observed an inverse U-shape relationship between green space exposure and self-rated health among elderly Chinese residents.^46^ A previous study examining the association between NDVI and birth weight demonstrated an inverted U-shaped relationship, indicating that birth weight was highest at moderate NDVI levels.^47^

In our study, we observed a U-shaped association between NDVI and the incidence of PE. In logistic regression analyses, compared with low residential greenness exposure, only moderate levels of exposure showed significant protective associations, whereas high NDVI exposure, although associated with lower incidence, did not reach statistical significance. These findings suggest that, while moderate levels of residential greenness may be effective in reducing the risk of PE compared to low levels, excessive greenness does not confer additional benefits and may even be counterproductive in some cases. This pattern aligns with findings demonstrating that the benefits of green space for physical activity do not increase linearly but rather plateau after reaching an optimal level.^48^ The U-shaped patterns were similar between urban and non-urban areas at NDVI values of 0.08 or higher; however, the trends appeared to diverge below this value. Nonetheless, owing to the wide CI in these ranges, interpretations should be made with caution. Further studies are needed to clarify whether the observed non-linear association reflects a true biological mechanism or is influenced by residual confounding and the composite nature of NDVI, which captures various types of greenness and vegetation simultaneously.

### Urban and Non-Urban Areas

A Chinese study^49^ suggested that, in urban areas, NDVI itself does not directly affect BP but exerts its effects through the mitigation of air pollution. In contrast, in non-urban areas, the mitigation of air pollution was only a partial factor, and NDVI directly influenced BP. Considering the low levels of air pollution in the region studied here (PM_2.5_ levels were 40.8 µg/m³ in the previous study and 10.9 µg/m³ in the present study), the lack of significant effects of residential greenness on air pollution mitigation could explain the absence of associations in urban areas observed in this study.

Meanwhile, in non-urban areas, the "direct effects" of residential greenness may have contributed to the lower incidence of PE. Further research is needed to determine what these "direct effects" are and why they are observed only in non-urban areas. Another study reported that the protective effects of greenness against oxidative stress, a major contributor to PE pathophysiology, also differed according to urbanization status.^50^ Collectively, these studies suggest that greenness may influence the pathophysiology of PE through different mechanisms in urban and non-urban areas. Potential hypotheses include differences in the quality of greenness between urban and non-urban areas,^51–53^ which may lead to variations in landscape characteristics and vegetation types.^54^ These factors, along with potential differences in accessibility to green spaces, could influence physical activity and antigenic exposures, ultimately impacting physiological responses.

### HDP Subtypes and Timing of PE Onset

In this study, an association was observed between residential greenness and PE, particularly LOPE. In contrast, no significant association was observed for GH, with odds ratios close to unity. This finding suggests that greenness may affect pregnancy-specific biological processes, including inflammation and immune regulation,^5,7,8^ beyond its potential effects on maternal blood pressure, possibly through pathways such as increased physical activity or reduced stress. This interpretation is supported by previous studies that reported beneficial effects of greenness on fetal growth indicators such as small for gestational age and low birth weight.^36^ Notably, EOPE, which is generally more severe and closely linked to placental pathology, exhibited a distinct pattern from other HDP subtypes. One possible explanation is that the U-shaped association observed in our study may have shifted to the left in pregnancies at high risk for EOPE, implying that the threshold at which greenness transitions from protective to potentially harmful may occur at a lower level of greenness exposure. This potential leftward shift raises the possibility of the effect of heterogeneity of greenness exposure, which should be considered and warrants further investigation in future studies. Moreover, given the small number of EOPE cases and wide CI, these findings should be interpreted with caution. For SP, although the association was not statistically significant, the direction of the association was protective and consistent with prior findings.^24,25^

### Clinical Implications

Although this was an observational study and causality could not be established, our findings may help inform decisions about the residential environment during pregnancy, such as choosing to live near areas with abundant surrounding greenery. Moreover, our study suggests that spending time in green spaces could be a simple and practical way to lower the risk of PE, especially in non-urban areas. While light physical activity is already encouraged in routine antenatal care, our findings highlight the potential additional benefits of engaging in such activities specifically in green environments— such as parks, forests, or tree-lined areas. Although the exact mechanisms underlying these benefits, such as stress reduction or effects on the immune system, are not fully understood, this study highlights the need to consider both environmental and individual factors to improve outcomes in pregnant women.

### Strengths and Limitations

This study investigated the association between residential greenness and PE by adjusting for a wide range of geographical factors, including air pollution, and individual-level factors such as smoking, alcohol consumption, physical activity, psychological distress, and social isolation. Furthermore, the inclusion of other HDP subtypes enabled a more detailed clinical interpretation. Notably, this is the first study on the association between residential greenness and PE in Japan.

This study has some limitations. First, as NDVI data were linked to participants by postal code, spatial inaccuracy in residential locations may have introduced nondifferential measurement errors, especially for smaller buffer sizes, which would bias effect estimates.^55^ Second, the study only covered Miyagi Prefecture and may not be generalizable to other regions, even within Japan, due to differences in vegetation types. Third, NDVI was used to quantify the degree of residential greenness, but factors such as accessibility to greenness and vegetation characteristics were not considered, which may limit the comprehensiveness of the analysis. Finally, the choice of residence may reflect pre-existing lifestyle habits, personality traits and socio-economic status, which may result in residual confounding in the observed associations.

## Conclusion

Moderate residential greenness was significantly associated with a lower incidence compared to low residential greenness. These results suggest that moderate residential greenery is worth considering when selecting where to live during pregnancy.

## Data Availability

Individual data are available upon request from the corresponding author after approval from the Ethical Committee and the Materials and Information Distribution Review Committee of the Tohoku Medical Megabank Organization.

## Acknowledgments

The authors would like to thank all the participants who consented to participate in this study and all the staff at the Tohoku Medical Megabank Organization, Tohoku University, Iwate Tohoku Medical Megabank Organization, and Iwate Medical University. A full list of the members of the Tohoku Medical Megabank Organization is available at https://www.megabank.tohoku.ac.jp/english/a240901/.

We used the artificial intelligence tool GPT-4o, developed by OpenAI, to assist in the initial drafting and editing of this manuscript. The tool provided suggestions for sentence structure and grammatical corrections. All intellectual contributions and final edits were made by the authors.

## Authors’ contributions

Conceptualization: Hisashi Ohseto, Ami Uematsu, Mami Ishikuro, Zheng Xian, Yuta Takahashi

Methodology: Hisashi Ohseto, Ami Uematsu, Mami Ishikuro, Zheng Xian, Yuta Takahashi

Visualization: Hisashi Ohseto

Supervision: Taku Obara, Tomoki Nakaya, Shinichi Kuriyama

Writing – original draft: Hisashi Ohseto

Writing – review & editing: All authors

## Information on the previous presentation

This work has not been previously presented anywhere.

